# A simple mathematical model for Coronavirus (COVID-19)

**DOI:** 10.1101/2020.04.23.20076919

**Authors:** Said Melliani, Abdelati El Allaoui, Lalla Saadia Chadli

## Abstract

A novel coronavirus (COVID-19) was identified in Wuhan, China in the end of 2019, it causing an outbreak of viral pneumonia. It caused to the death rate of 4.63% among 571, 678 confirmed cases around the world to the March 28th, 2020. In this brief currentstudy, we will present a simple mathematical model where we show how the probability of successfully getting infected when coming into contact with an infected individual and the per-capita contact rate affect the healthy and infected population with time. The proposed model is used to offer predictions about the behavior of COVID-19 for a shorter period of time.

## 1 Introduction

Early January 2020, a novel coronavirus (COVID-19) was identified as the infectious agent causing an outbreak of viral pneumonia in Wuhan, China, despite, the first cases had their symptom onset in December 2019. It has caused at least 571, 678 confirmed cases with 26, 494 deaths and 158, 700 recovered in the world, from the start of this pandemic until March 28th, 2020, according to World Health Organization [2]. The ongoing outbreak of coronavirus disease is affecting 199 countries. The most infected countries distributed in descending order of infected cases are: USA, Italy, Spain, China, Germany, Iran and France and other less infected countries. The seriousness of this epidemic is evident in the speed of its spread with a proportion of 4.63% of deaths among the infected worldwide, which is a low proportion compared to those of other epedemics.

Since the beginning of COVID-19, Several researchers attacked the study of this pandemic, each of his field [5, 13–15]. Also, Mathematicians contributed their part to these studies. In [10], Qianying Lin et al studied the transmission of COVID-19 in Wuhan, China with more details. In [11], Chen et al. treated a mathematical model for simulating the phase-based transmissibility of a novel coronavirus, the goal of this study was to calculate the transmissibility of the virus by using a developed mathematical model. Other authors have studied this new disease mathematically (see [5, 11, 15]).

The purpose of this paper is to consider a mathematical model to study predictions of the outbreak of coronavirus in the most infected countries in the world such as: USA, Italy, Spain, Germany, Iran and France (China is excluded from these studies because several works have been devoted only to China [4–6,10,12–15]). And, how to reduce the seriousness of Coronavirus which is the speed of its outbreak. For that, based on [1], we consider the following system of non-linear differential equations:

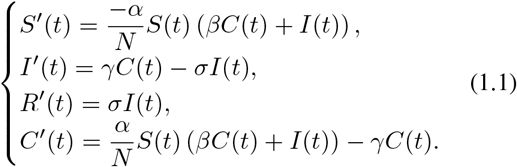

Where the meaning of the parameters is given as follows:

*S*(*t*) the susceptible at time *t, I*(*t*) the infectious at time *t, R*(*t*) the removed at time *t, C*(*t*) the exposed at time *t*,

*N* the population studied (total population), *α* is an average of the number of infected people by contact with one symptomatic individual who has been infected with Corona virus,

*γ* is the per-capita infectious rate,

*σ* is the per-capita death rate.

We shall assume that the population *N* is constant, and *N* = *S*(*t*) + *E*(*t*) + *I*(*t*) + *R*(*t*). we are interested in predictions of the outbreak of the novel coronavirus disease (COVID-19) for the next 15 weeks from March 29th, 2020. At the same time, we are interested in how the spread of this infectious disease. We have followed the Corona disease outbreak news from the start of this pandemic, January 10th to March 28th, 2020.

The table 1 shows the numbers of confirmed cases of Corona virus for the most infected countries in the world:

**Table 1:** Numbers of confirmed cases of COVID-19 outbreak as they appear in official data obtained from WHO from January 10th to March 28th, 2020.

| Country | Total infected cases | Total Deaths | Total recovered | Mortality rate |
| --- | --- | --- | --- | --- |
| USA | 120,076 | 1,993 | 3,229 | 1.65% |
| Italy | 92,472 | 10,023 | 12,384 | 10.83% |
| Spain | 72,335 | 5,820 | 12,285 | 8.04% |
| Germany | 57,695 | 430 | 8,481 | 0.74% |
| France | 37,575 | 2314 | 5,700 | 6.15% |
| Iran | 35,408 | 2,517 | 11,679 | 7.10% |

In this paper, we will consider the following parameters:

Coronavirus is twice as contagious as the flu. Research signals a person with the flu infects an average of 1.28 other people, CNN Chief Medical Correspondent Dr. Sanjay Gupta said. But with coronavirus, “it’s likely between two and three” other people”. For these reasons and based on the paper [10], we suggest that transmission rate *α* varies between 0.5944 and 1.68. Coronavirus can be transmitted by people who are infected by this pandemic. The chance of getting infected for an infectious individual is very higher. which means that the factor *β* satisfies 0 *≤ β ≤* 1.

In the following figures we will show the prediction of our model given by the proposed system of non linear differential equations in a shorter interval of time [0 15], where 0 is the day the predictions start, it’s March 29th, 2020, and 15 designates the 15th week, i.e. from March 29th to July 11th, 2020.

Figures 1.1, 1.2, 1.3, 1.4, 1.5 and 1.6 show cumulative predictions of infected in USA, Italy, Spain, Germany, France and Iran respectively from March 29th to July 11th, 2020. Where we assumed that: *α* = 0.6 and *β* = 0.037, with the other parameters given in the table 2. To analyze these figures, for example from figure 1.1, we can notice that the growth of the infected is faster than the other countries. We can see that the predictions of the number of infected with COVID-19 towards the end of the 15th week may reach: 1.944 *×* 10^6^ cases in USA, 8.1168 *×* 10^5^ in Italy, 8.1144 *×* 10^5^ in Spain, 8.825 *×* 10 in Germany, 4.4216 *×* 10 in France and 3.979 *×* 10^5^ in Iran.

**Table 2:** Summary table of the parameters used in model (1.1)

| Country | Population N | S(0) Assumed $0.8 \times N$ | I(0) | R(0) | C(0) Assumed | $\sigma$ | $\gamma = \frac{I(0)}{C(0)}$ |
| --- | --- | --- | --- | --- | --- | --- | --- |
| USA | 330,505,054 | 264,404,043 | 120,076 | 5,222 | 65,975,713 | 0.0165 | 0.0018 |
| Italy | 60,484,425 | 48,387,540 | 92472 | 22371 | 11982042 | 0.1083 | 0.0077 |
| Spain | 46,750,165 | 37,400,132 | 72,335 | 18,105 | 9,259,593 | 0.0804 | 0.0078 |
| Germany | 83,715,478 | 66,972,382 | 57,695 | 8,911 | 16,676,490 | 0.0074 | 0.0034 |
| France | 65,236,642 | 52,189,314 | 37,575 | 8,014 | 13,001,739 | 0.0615 | 0.0028 |
| Iran | 83,715,157 | 66,972,126 | 35,408 | 14,196 | 16,693,427 | 0.0710 | 0.0021 |

**Figure 1.1:**
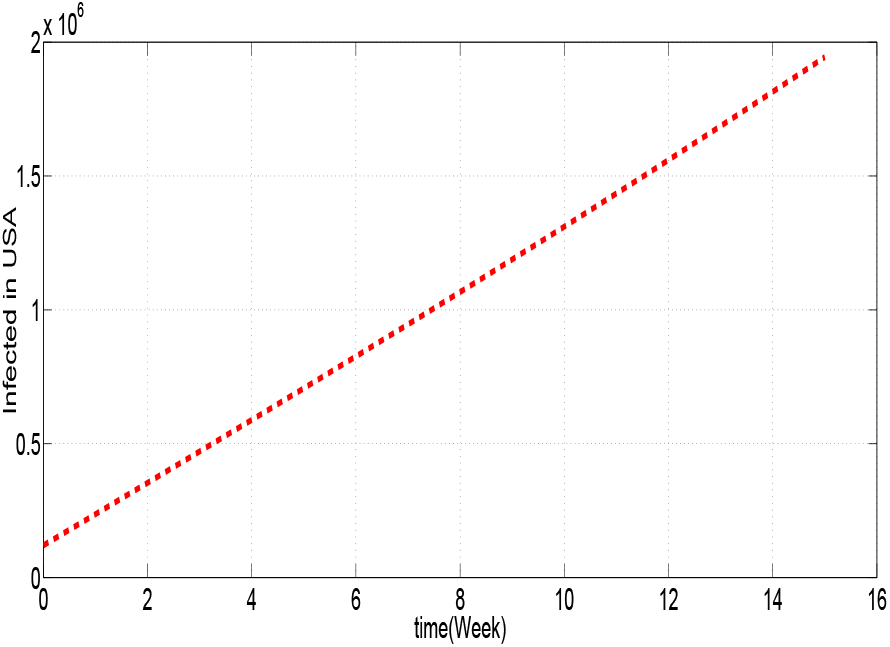
Model prediction for USA.

**Figure 1.2:**
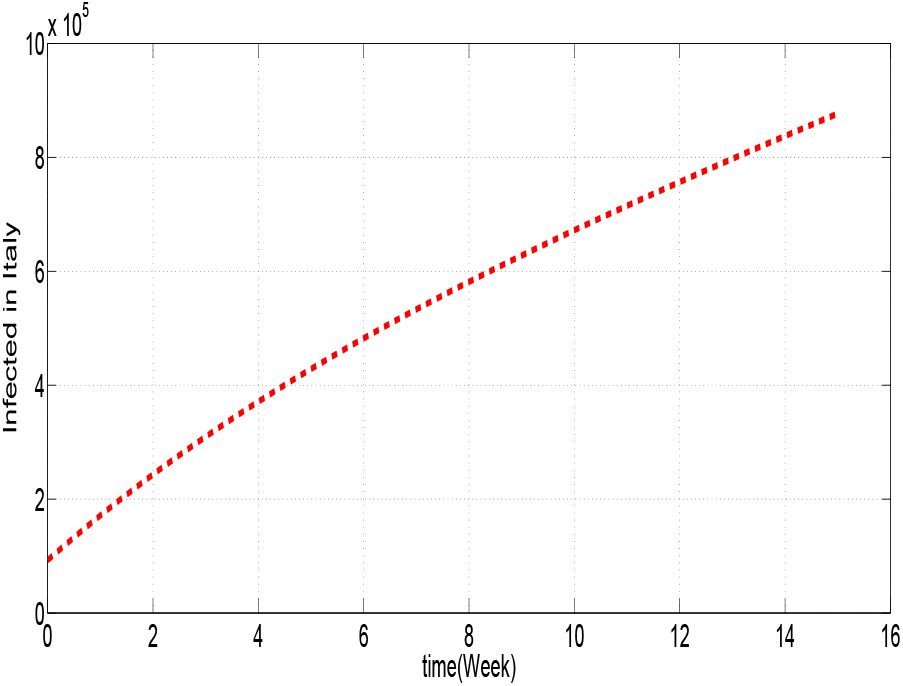
Model prediction for Italy.

**Figure 1.3:**
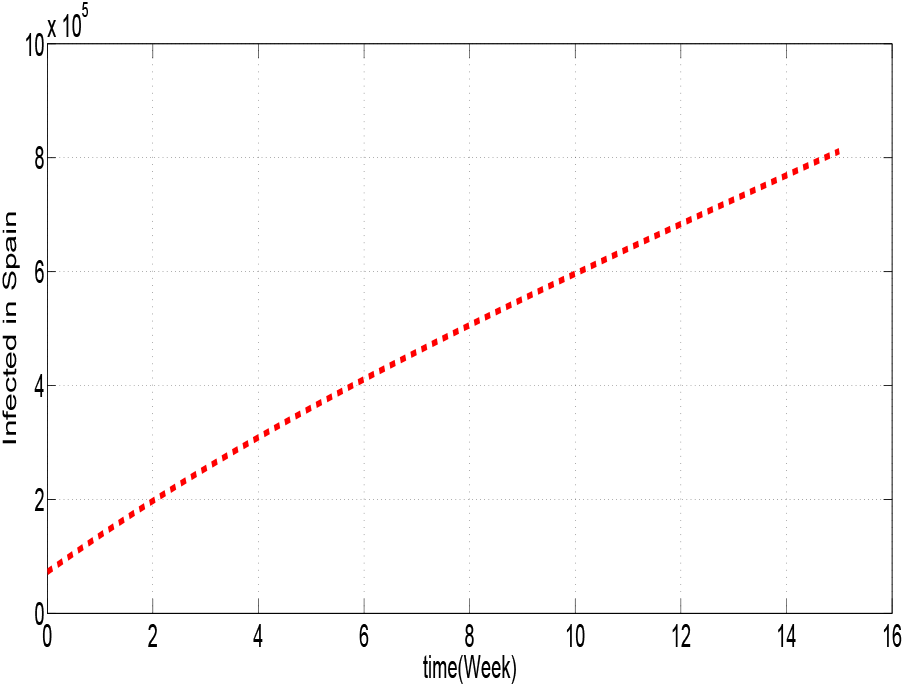
Model prediction for Spain.

**Figure 1.4:**
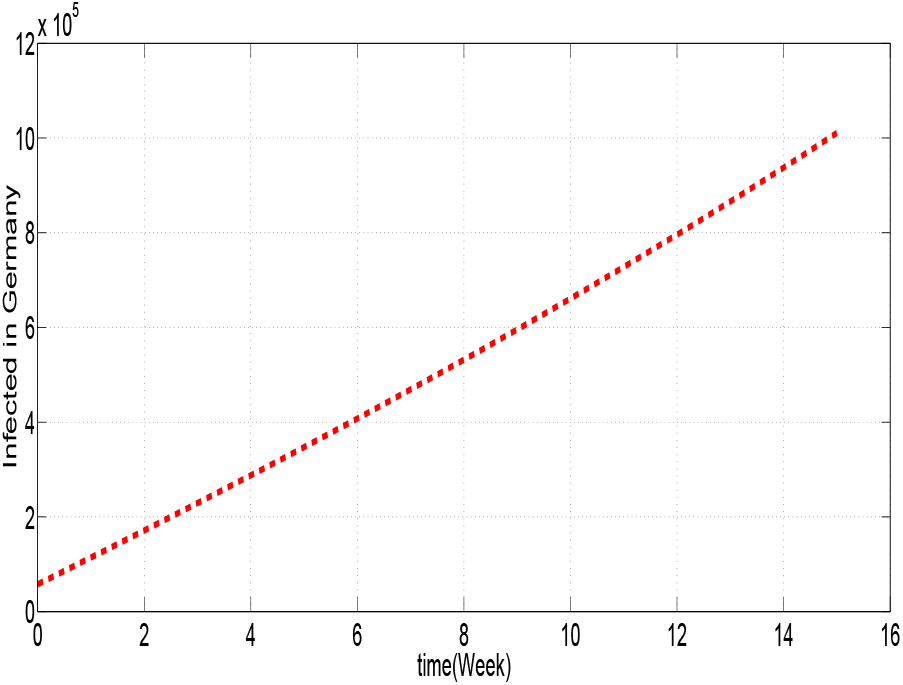
Model prediction for Germany.

**Figure 1.5:**
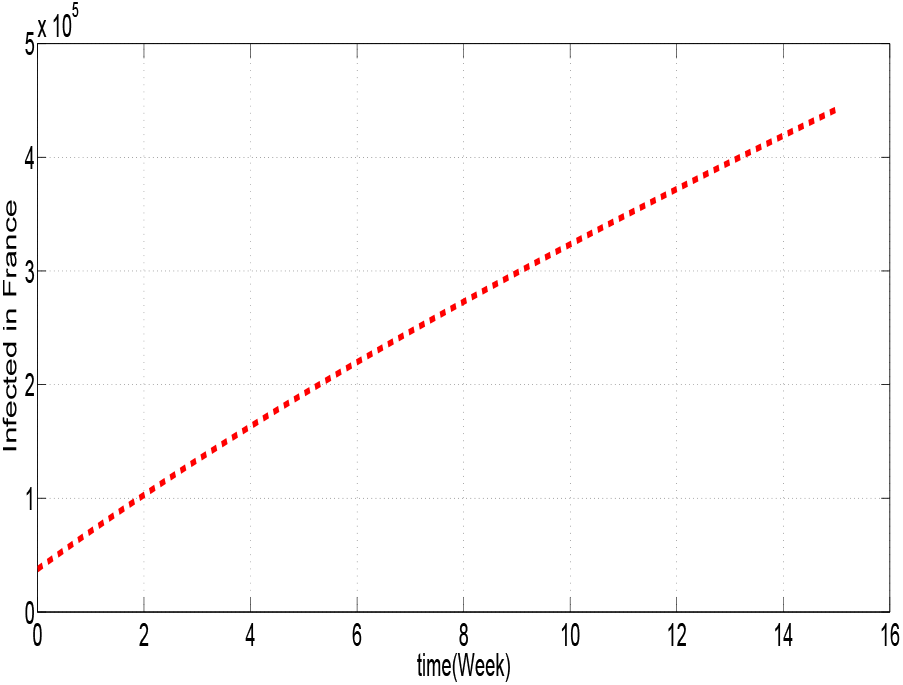
Model prediction for France.

**Figure 1.6:**
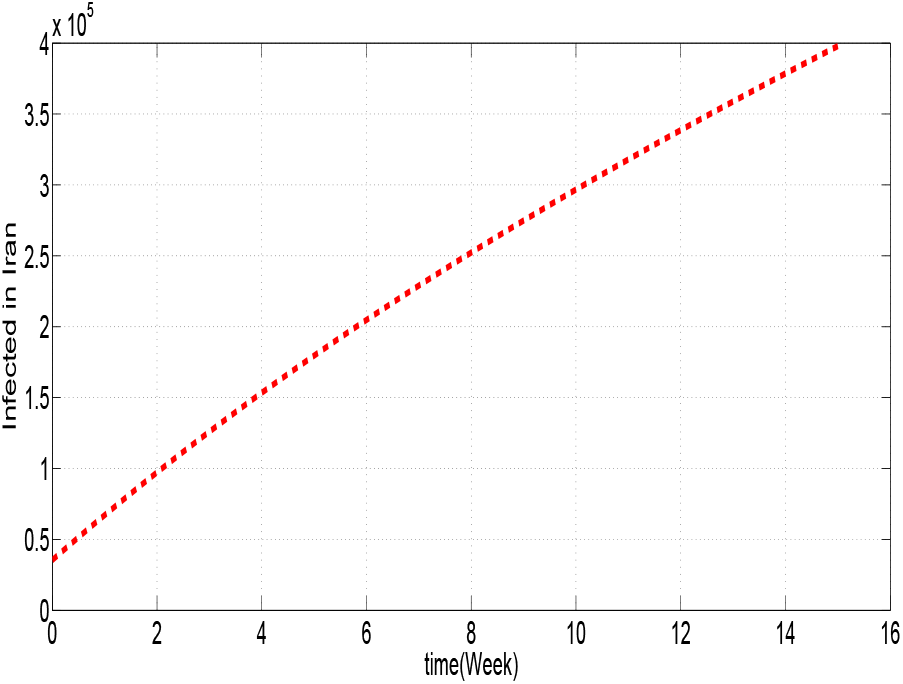
Model prediction for Iran.

In the following figures, we will present the number of infected for USA and Italy most infected in the world for different values of *α*:

From figure 1.7, we can see that, the higher the coefficient, the higher the number of infected, likewise for the case of Italy in figure 1.8. So to have a minimum possible of infected, we must have a minimum value of *α*. Since, the parameter is defined by the product of the probability of successfully getting infected when coming into contact with an infected individual and the per-capita contact rate, for that, it is necessary to decrease these two parameters which depend both on “the contact”. That is why reducing contact remains among the reasons for reducing infections.

**Figure 1.7:**
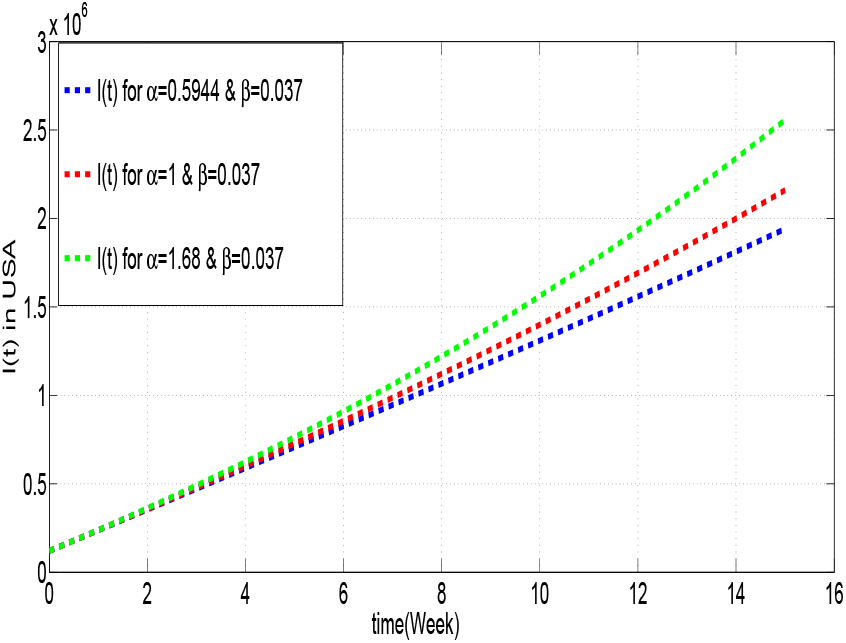
Model prediction for USA with different values of *α*.

**Figure 1.8:**
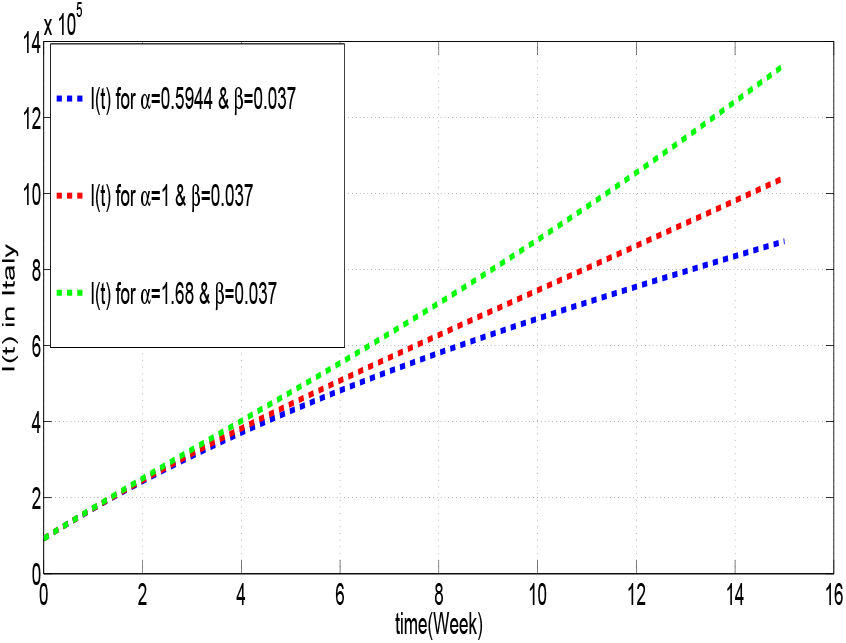
Model prediction for Italy with different values of *α*.

To evaluate the equilibrium points, we solve the following equations: *S*′(*t*) = *I*′(*t*) = *R*′(*t*) = *C*′(*t*) = 0.

That is,

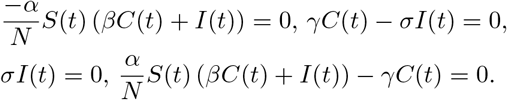

Which is equivalent to *I*(*t*) = *C*(*t*) = 0.

The Jacobian is given by

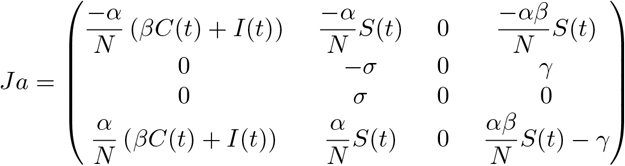

The Jacobian Calculated at the equilibrium point, i.e. we take into account *I*(*t*) = *C*(*t*) = 0 will be given by

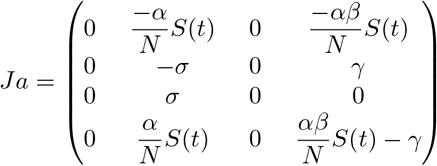

Whose eigenvalues are given by

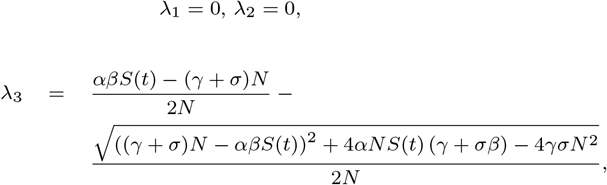

and

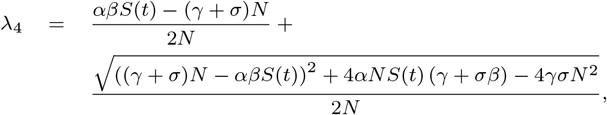

since, the parameters of the studied model verify

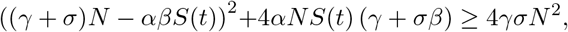

For the case of USA, we get

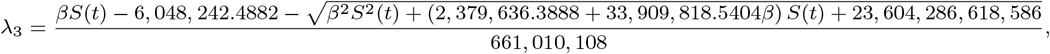

and

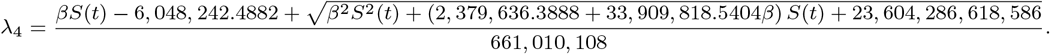

With small calculations, we can easily verify that

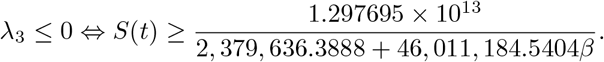

We have *β ∈* [0 1], so, a sufficient condition for *λ*_3_ to be negative is *S*(0) *≥* 5, 453, 335, while in this case *λ*_4_ *≥* 0.

## Data Availability

Te data used to support the findings of this study
are available from the corresponding author upon
request

https://www.researchgate.net/profile/Abdelati_El_Allaoui2

## Data Availability

Te data used to support the findings of this study are available from the corresponding author upon request.

## Author’s contributions

All authors contributed equally to the writing of this paper. All authors read and approved the final manuscript.

## Acknowledgements

The authors express their sincere thanks to the anonymous referees for numerous helpful and constructive suggestions which have improved the manuscript.

